# Multimodal Deep Learning for ARDS Detection

**DOI:** 10.1101/2025.08.08.25333333

**Authors:** Stefan Broecker, Jason Y. Adams, Girish Kumar, Rachael A. Callcut, Yuan Ni, Thomas Strohmer

## Abstract

**Objective:** Poor outcomes in acute respiratory distress syndrome (ARDS) can be alleviated with tools that support early diagnosis. Current machine learning methods for detecting ARDS do not take full advantage of the multimodality of ARDS pathophysiology. We developed a multimodal deep learning model that uses imaging data, continuously collected ventilation data, and tabular data derived from a patient’s electronic health record (EHR) to make ARDS predictions.

**Materials and Methods:** A chest radiograph (x-ray), at least two hours of ventilator waveform (VWD) data within the first 24 hours of intubation, and EHR-derived tabular data were used from 220 patients admitted to the ICU to train a deep learning model. The model uses pretrained encoders for the x-rays and ventilation data and trains a feature extractor on tabular data. Encoded features for a patient are combined to make a single ARDS prediction. Ablation studies for each modality assessed their effect on the model’s predictive capability.

**Results:** The trimodal model achieved an area under the receiver operator curve (AUROC) of 0.86 with a 95% confidence interval of 0.01. This was a statistically significant improvement (p<0.05) over single modality models and bimodal models trained on VWD+tabular and VWD+x-ray data.

**Discussion and Conclusion:** Our results demonstrate the potential utility of using deep learning to address complex conditions with heterogeneous data. More work is needed to determine the additive effect of modalities on ARDS detection. Our framework can serve as a blueprint for building performant multimodal deep learning models for conditions with small, heterogeneous datasets.

## Introduction

Acute respiratory distress syndrome (ARDS) is a severe form of hypoxemic respiratory failure that has proven challenging to diagnose and manage in a timely fashion, despite affecting up to 10% of ICU admissions and 25% of mechanically ventilated (MV) patients [1]. ARDS is associated with substantial morbidity and mortality, prolonged MV, high hospital-associated costs, and long-term physical and psychologic dysfunctions [1, 2]. Moreover, poor outcomes in ARDS are associated with delayed and missed diagnosis and suboptimal use of evidence-based therapies, even by subspecialty-trained clinicians [1, 2, 3, 4], highlighting the need for tools to support early diagnosis.

The diagnostic criteria for ARDS require laboratory-based, electronic health record (EHR)-derived arterial blood gas (ABG) measurement of the degree of hypoxemia, ascertainment of mechanical ventilation settings, and the presence of bilateral opacities on chest imaging [5]. This inherent multimodality of ARDS pathophysiology suggests that integrating multiple data sources could improve early diagnostic accuracy. Recent advances in deep learning have shown promise for early ARDS detection using single modalities, including convolutional neural networks (CNNs) on single chest radiographs (CXRs) [6] and on ventilator waveform data (VWD) [7]. While these unimodal approaches have shown promise, they may miss important clinical patterns that only become apparent when analyzing multiple data sources simultaneously.

Previous work has explored bimodal approaches to ARDS detection through methods that combine the predictions of models trained on CXR and EHR data [8, 9, 10]. However, recent advances in multimodal machine learning outside of ARDS detection suggest that more integrated approaches to modality fusion of more complex data could yield better results. For instance, studies have demonstrated the utility of deep fusion techniques by using medical images and diagnostic notes to improve classification tasks including pneumonia detection, COVID detection, and bone abnormality detection [11]. These more sophisticated methods can capture both the unique contributions of each data type and the potentially important interactions between different physiologic measurements.

Recent work is also often limited to only two datatypes or modalities [10]. While bimodal approaches have shown initial promise, they have been largely limited to static types of data, such as a single metric of hypoxemia from arterial blood gases. This narrow use of data fails to account for the dynamic nature of patients with need for mechanical ventilation leading to missed opportunities to identify the earliest patterns of disease. The complex pathophysiology of ARDS, as suggested by the diverse modalities used in the Berlin criteria, suggests that integrating VWD, CXRs, and clinical data from the EHR could capture subtle interactions between physiologic, radiographic, and clinical manifestations of the condition.

Finally, past work in ARDS detection uses thousands to tens of thousands of patients’ data during training [10]. As collecting data from this volume of patients can be both expensive and time consuming, robust methods that can be trained with less data are needed.

Building on advances in both ARDS detection and multimodal learning, we hypothesized that combining VWD, CXRs, and EHR data into a single, trimodal deep learning model would improve both the accuracy and robustness of early ARDS detection, even when training on small, heterogeneous datasets. Specifically, we investigated whether an integrated trimodal model that uses pretrained encoders could better discriminate between ARDS and non-ARDS patients compared to unimodal and bimodal methods.

## Methods

### Cohort Description

All patient data were obtained as part of a prospective, Institutional Review Board approved study collecting data from mechanically ventilated adults admitted to the intensive care unit (ICU) at the University of California (UC) Davis Medical Center. Three critical care physicians performed retrospective chart review to identify the cause of respiratory failure in subjects from the study cohort enrolled between 2015 and 2019. The presence or absence of ARDS was determined using the Berlin consensus criteria [5] by independent clinician review, with disagreements between any two clinicians resolved by consensus chart review. Subjects with ARDS were divided into two subgroups as a function of relative ambiguity of ARDS classification: Group 1) patients with confirmed moderate or severe ARDS without chronic obstructive pulmonary disease and/or asthma (to minimize the risk of misclassifying ARDS as a result of concurrent non-ARDS acute or nonacute chronic lung disease-associated hypoxemia), chronic congestive heart failure, or severe obesity and Group 2) patients with any severity of ARDS and without exclusion of these conditions.

Causes of ARDS for patients meeting the Berlin criteria are shown in Table 1. Table 1 also includes basic demographic information and additional clinical information such as Sequential Organ Failure Assessment score, hospital length of stay, and hospital mortality.

**Table 1.** Comparison of characteristics between ARDS and non-ARDS patients. Note: 3 non-ARDS patients were not admitted to the ICU.

| Characteristic | ARDS (n = 120) | Non-ARDS (n = 120) |
| --- | --- | --- |
| Age (median [IQR]) | 57 (47-68) | 62 (53-71) |
| Female (n [%]) | 53 (44) | 48 (40) |
| Body mass index (median [IQR]) | 27.5 (22.4-34.7) | 26.6 (22.2-33) |
| Sequential Organ Failure Assessment score <sup>1</sup>(median [IQR]) | 11 (9-14.3) | 8 (6-11) |
| Days from intubation to Berlin criteria (median [IQR]) | 0.11 (0.06-0.24) | — |
| Median PaO<sub>2</sub>/FiO<sub>2</sub> first 24 hours (median [IQR]) | 166 (132-219.5) | 311 (218-390) |
| Worst PaO<sub>2</sub>/FiO<sub>2</sub> first 24 hours (median [IQR]) | 111 (73-153) | 218 (135-313) |
| Hospital length of stay (median [IQR]) | 12.2 (7.3-22.7) | 9 (5.3-16.9) |
| Hospital mortality (n [%]) | 57 (48) | 32 (27) |
| Ventilator-free days in 28 days (median [IQR]) | 6.6 (0-21.9) | 23.5 (4-26.3) |
| **ARDS insult type (n [%])** |  |  |
| Pneumonia | 64 (53.3) | — |
| Aspiration | 31 (25.8) | — |
| Non-pulmonary sepsis | 18 (15) | — |
| Trauma | 3 (2.5) | — |
| Other | 4 (3.3) | — |

No sample size calculation was conducted for this study; however, sample size was guided by the range of cohort sizes in previous studies of VWD analysis [7] and to achieve a balanced dataset for machine learning (ML) model development. Standard ML algorithms are often biased towards the majority class, resulting in a higher misclassification rate for the minority class [12].

### Data Preparation

VWD representing air flow in liters per minute was collected from Puritan Bennett 840 ventilators as a 1-dimenional time series at a rate of 50Hz. VWD for each patient was split into windows of 30,000 samples (10 minutes of data collection), and the number of windows for each patient was limited to the first 36 available windows (the first six hours of available data). Incomplete windows were padded with 0s to create uniform window lengths. All patients had at least one hour of VWD data in the first 24 hours after Berlin criteria were first met or after the start of MV.

CXRs were obtained from a Picture Archiving and Communication System (PACS) in the form of DICOM files. For ARDS patients, the first CXR that met the Berlin criteria for ARDS was used. For non-ARDS patients, the first CXR obtained within 24 hours of intubation was used. CXRs were resized and cropped to a uniform size of 512×512 pixels.

Tabular data derived from the EHR were included from the first 24 hours after meeting Berlin criteria or the first 24 hours after the start of MV for non-ARDS patients. A full list of features used is available in Supplementary Table 1. Categorical features were one-hot encoded during data processing. Numerical features that were measured multiple times for a single patient were aggregated. For each numerical feature, we calculated the minimum, maximum, mean, and median of the feature. All numerical features were then normalized to have a mean of zero and unit variance. Missing numerical features were filled with the mean of the feature across the dataset.

During model training, each VWD window was treated as a separate training instance, with the corresponding CXR and EHR data repeated across windows for the same patient. During inference, predictions from all windows for a single patient were averaged to produce a final classification probability.

### Train/Test Splitting

All data splits were performed at the patient level. A 5-fold k-fold split procedure (outer folds) was used, with a further nested 5-fold split (inner folds) on each outer fold. Hyperparameter tuning was performed using a grid search on inner folds. Within each training split, data was further split 80/20 into training and validation splits. The splitting procedure can be seen in Supplementary Figure 3.

The entire k-fold procedure was repeated 10 times for each modality combination, and performance was assessed using unseen data from the outer k-fold splits. This resulted in 50 total holdout sets for each model. One-hot encoding and scaling, as described in the previous section, were calculated on the training set for each split and applied to the validation and holdout sets.

### Model Development

Each multimodal model consists of sub-classes of models: base encoders for each modality, modality-specific projection heads, and a classification head. Given a batch of patients, the multimodal model first encodes data for each patient from each modality using the base encoders. Then it passes the encoded data through modality-specific projection heads. Finally, it combines the projected data via concatenation before passing it into the single classification head. Figure 1 shows this graphically.

**Figure 1.**
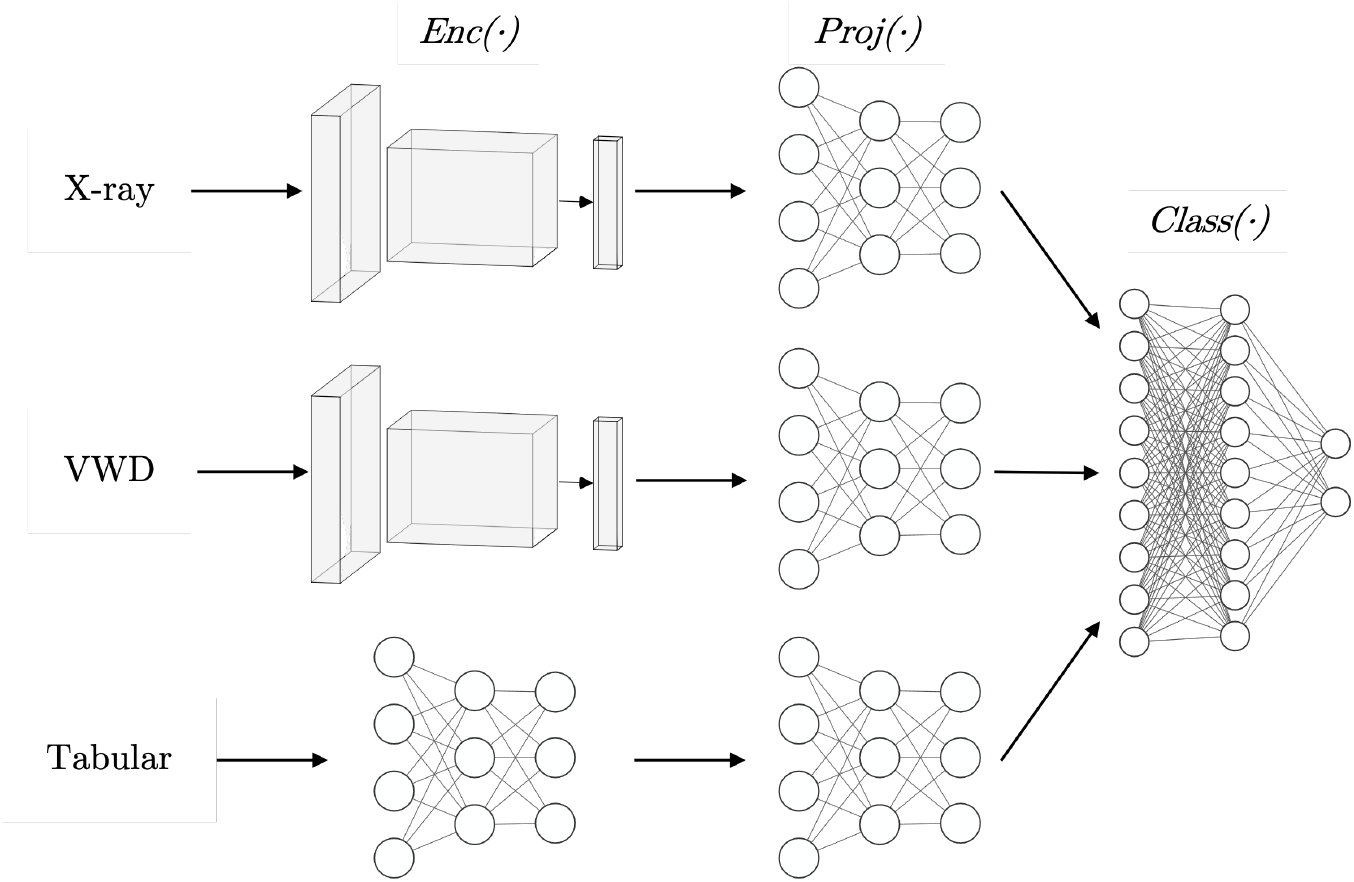
A schematic depiction of our trimodal architecture showing encoders, projection heads, and classification layers. X-ray and ventilator waveform encoders use a convolutional neural network architecture, while tabular encoders, projection heads, and classification networks use fully connected layers.

Modality-specific base encoders, *Enc*(·), map an input sample, **x**, to an embedding vector, 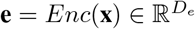, where *D*_*e*_ is the embedding dimension size. *D*_*e*_ is 128 for both CXR and VWD data, and 64 for EHR data. Base encoders are described in detail in the next section.

Projection heads, *Proj*(·), map **e** to vector 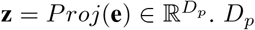 is a tuned as a hyperparameter for each model, with possible values of 8, 16, or 32. *Proj*(*·*) is a one- or two-layer perceptron network with hidden layer and output layer dimensions of *D*_*p*_. **z** is normalized to the unit hypersphere in 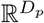. The number of layers in *Proj*(·) is tuned as a hyperparameter.

Classification heads, *Class*(·), map **z** to two classes representing ARDS or non-ARDS. *Class*(·) is a two-layer perceptron network with input and hidden layer dimension of *D*_*p*_*∗m*, where *m* is the number of modalities, and output layer dimension of two with a softmax activation.

*Enc*(·) layers are pretrained models (CXR and VWD) or trained on each individual split (EHR). *Enc*(·) layers are frozen during downstream model training. *Proj*(·) and *Class*(·) layers are jointly trained using a cross-entropy loss.

Models are trained using the Adam optimizer with an initial learning rate of 1e-4 and weight decay of 1e-6. Training is performed for a maximum of 150 epochs with early stopping based on validation loss with a patience of 3 epochs after a warm-up of 10 epochs.

### Base Encoders

The VWD-specific encoder is a CNN pretrained for audio tagging, the task of assigning one or multiple semantic labels to an audio clip [13]. We use the pretrained model to extract intermediate features to use as our embeddings [14].

The CXR-specific encoder uses a pretrained model, which is a ResNet-50 backbone trained on CXRs for eight cardiopulmonary radiological tasks [15, 16]. We remove the final projection head of the model and use the remaining network to generate CXR embeddings.

The tabular-specific encoder is a model pretrained using the VIME (Value Imputation and Mask Estimation) framework [17]. This approach trains two networks on tabular data corrupted by data masks: one for data reconstruction and another for mask estimation. We use an intermediate network that serves as input to the two networks as our encoder for generating tabular embeddings.

### Performance Assessment

We investigated three individual unimodal models and four combined multimodal models. The four combined multimodal models were: VWD+CXR, VWD+tabular, CXR+tabular, and trimodal.

Final model selection for each modality combination was based on the area under the receiver operating characteristic curve (AUROC) as measured on the validation set. Model performance on unseen data from the holdout datasets was assessed by the AUROC, area under the precision-recall curve (AUPRC) and typical confusion matrix statistics. Performance metrics are reported with 95% confidence intervals calculated with python’s SciPy library [18]. Hypothesis testing was performed between each modality combination with two-sample t-tests. Full p-values are in Supplementary Table 5. Model calibration was assessed descriptively using calibration curves plotting the predicted probability of ARDS against the observed prevalence of ARDS across predicted probability strata.

## Results

### Overall Model Performance

Across all evaluated modality combinations, the trimodal model achieved the highest overall performance with an AUROC of 0.86 (CI: 0.01) and an overall accuracy of 78% (CI: 0.02) when averaged across all holdout datasets. The trimodal model achieved a statistically significant improvement over all unimodal models, as well as the VWD+Tabular and VWD+CXR bimodal models (p < 0.05) but not the CXR+Tablular model. Table 2 shows the full complement of discrimination metrics averaged across all holdout datasets for models trained on each modality combination.

**Table 2.** Average metrics for models trained on each modality combination. Best values for each metric and cohort are shown in bold.

| Modality | Cohort | Accuracy | AUROC | Precision | Sensitivity | Specificity | AUPRC |
| --- | --- | --- | --- | --- | --- | --- | --- |
| Tabular | All | 0.77 ± 0.01 | 0.83 ± 0.01 | **0.78** ± 0.01 | 0.74 ± 0.03 | **0.79** ± 0.02 | 0.82 ± 0.02 |
| X-ray | All | 0.69 ± 0.02 | 0.76 ± 0.02 | 0.68 ± 0.04 | 0.69 ± 0.04 | 0.68 ± 0.04 | 0.78 ± 0.02 |
| VWD | All | 0.71 ± 0.02 | 0.78 ± 0.02 | 0.72 ± 0.02 | 0.71 ± 0.03 | 0.72 ± 0.03 | 0.79 ± 0.02 |
| VWD+X-ray | All | 0.73 ± 0.01 | 0.82 ± 0.02 | 0.74 ± 0.02 | 0.72 ± 0.03 | 0.74 ± 0.02 | 0.84 ± 0.01 |
| VWD+Tabular | All | 0.76 ± 0.02 | 0.84 ± 0.02 | 0.76 ± 0.02 | 0.77 ± 0.03 | 0.75 ± 0.03 | 0.83 ± 0.02 |
| X-ray+Tabular | All | 0.76 ± 0.02 | 0.85 ± 0.02 | 0.78 ± 0.02 | 0.75 ± 0.03 | 0.78 ± 0.03 | 0.86 ± 0.02 |
| Trimodal | All | **0.78** ± 0.02 | **0.86** ± 0.01 | **0.78** ± 0.02 | **0.78** ± 0.03 | 0.78 ± 0.03 | **0.87** ± 0.02 |

### Modality Contribution Analysis

In two of the three unimodal models, we observed consistent performance gains across all measures of discrimination when adding either a second or third modality, with the trimodal model showing the highest overall performance (Table 2). The model combining VWD+CXR data yielded substantially better performance than either modality alone (VWD+CXR model AUROC of 0.82 (CI: 0.02), compared to unimodal VWD and CXR models with AUROCs of 0.78 (CI: 0.02) and 0.76 (CI: 0.02), respectively. In contrast, bimodal models that used Tabular data from the EHR did not consistently outperform the unimodal model using Tabular data alone, with bimodal improvement in AUROC, AUPRC, and sensitivity but not in other metrics.

### Performance Stratification by Patient Group

Model performance varied notably between Group 1 (more clear-cut ARDS cases) and Group 2 (less clear-cut cases). In Group 1, the trimodal model achieved the highest AUROCs of all models for both Group 1 (0.96, CI: 0.01) and Group 2 0.78 (CI: 0.02) ARDS cases. We observed a consistent performance gap between Group 1 and Group 2 ARDS cases across all modalities, but the gap was not uniform (Table 3). For unimodal models, VWD data showed the largest performance change (ΔAUROC = 0.23), followed by x-ray data (ΔAUROC = 0.18), and then tabular data (ΔAUROC= 0.17). In bimodal models, VWD+Tabular and CXR+Tabular had the same performance decrease (ΔAUROC = 0.18), while the VWD+ CXR model had a larger drop (ΔAUROC = 0.20

**Table 3.** Average metrics for models trained on each modality combination. Best values for each metric and cohort are shown in bold.

| Modality | Cohort | Accuracy | AUROC | Precision | Sensitivity | Specificity | AUPRC |
| --- | --- | --- | --- | --- | --- | --- | --- |
| Tabular | Group 1 | $0.85 \pm 0.02$ | $0.93 \pm 0.02$ | $0.86 \pm 0.04$ | $0.82 \pm 0.04$ | $0.88 \pm 0.03$ | $0.92 \pm 0.02$ |
| X-ray | | $0.79 \pm 0.03$ | $0.86 \pm 0.03$ | $0.79 \pm 0.05$ | $0.72 \pm 0.05$ | $0.84 \pm 0.03$ | $0.88 \pm 0.03$ |
| VWD | | $0.82 \pm 0.02$ | $0.90 \pm 0.02$ | $0.82 \pm 0.03$ | $0.83 \pm 0.03$ | $0.81 \pm 0.04$ | $0.91 \pm 0.02$ |
| VWD+X-ray | | $0.85 \pm 0.02$ | $0.92 \pm 0.02$ | $0.87 \pm 0.03$ | $0.81 \pm 0.03$ | $0.88 \pm 0.03$ | $0.93 \pm 0.02$ |
| VWD+Tabular | | $0.85 \pm 0.03$ | $0.93 \pm 0.02$ | $0.85 \pm 0.03$ | **$0.85 \pm 0.04$** | $0.85 \pm 0.03$ | $0.93 \pm 0.02$ |
| X-ray+Tabular | | $0.86 \pm 0.02$ | $0.94 \pm 0.01$ | $0.88 \pm 0.03$ | $0.83 \pm 0.03$ | $0.89 \pm 0.03$ | $0.94 \pm 0.02$ |
| Trimodal | | **$0.88 \pm 0.02$** | **$0.96 \pm 0.01$** | **$0.91 \pm 0.03$** | $0.84 \pm 0.04$ | **$0.92 \pm 0.03$** | **$0.96 \pm 0.01$** |
| Tabular | Group 2 | **$0.70 \pm 0.02$** | $0.76 \pm 0.02$ | **$0.73 \pm 0.03$** | $0.67 \pm 0.04$ | **$0.73 \pm 0.03$** | $0.76 \pm 0.03$ |
| X-ray | | $0.62 \pm 0.02$ | $0.68 \pm 0.03$ | $0.61 \pm 0.04$ | $0.67 \pm 0.04$ | $0.56 \pm 0.05$ | $0.74 \pm 0.02$ |
| VWD | | $0.63 \pm 0.03$ | $0.67 \pm 0.03$ | $0.65 \pm 0.03$ | $0.62 \pm 0.04$ | $0.64 \pm 0.04$ | $0.71 \pm 0.03$ |
| VWD+X-ray | | $0.65 \pm 0.02$ | $0.72 \pm 0.03$ | $0.66 \pm 0.03$ | $0.66 \pm 0.04$ | $0.64 \pm 0.03$ | $0.76 \pm 0.03$ |
| VWD+Tabular | | $0.69 \pm 0.02$ | $0.75 \pm 0.03$ | $0.71 \pm 0.03$ | $0.72 \pm 0.04$ | $0.68 \pm 0.04$ | $0.76 \pm 0.03$ |
| X-ray+Tabular | | $0.69 \pm 0.02$ | $0.76 \pm 0.02$ | $0.72 \pm 0.03$ | $0.70 \pm 0.04$ | $0.69 \pm 0.04$ | **$0.80 \pm 0.03$** |
| Trimodal | | **$0.70 \pm 0.02$** | **$0.78 \pm 0.02$** | $0.71 \pm 0.03$ | **$0.73 \pm 0.03$** | $0.68 \pm 0.04$ | **$0.80 \pm 0.02$** |

### Model Calibration

The trimodal model was generally well-calibrated across its range of predicted probabilities in the holdout datasets, with a small overprediction of ARDS risk toward the lower range of predictions and similarly small underprediction of risk in the upper range (Figure 2). Supplementary Table 6 shows the performance of the trimodal model as assessed by confusion matrix statistics across deciles of model classification thresholds. Note that the number of patients in each decile of predicted probability was not uniform and models made no predictions with probability < 0.2 or > 0.8 (grey histogram bars, Figure 2).

**Figure 2.**
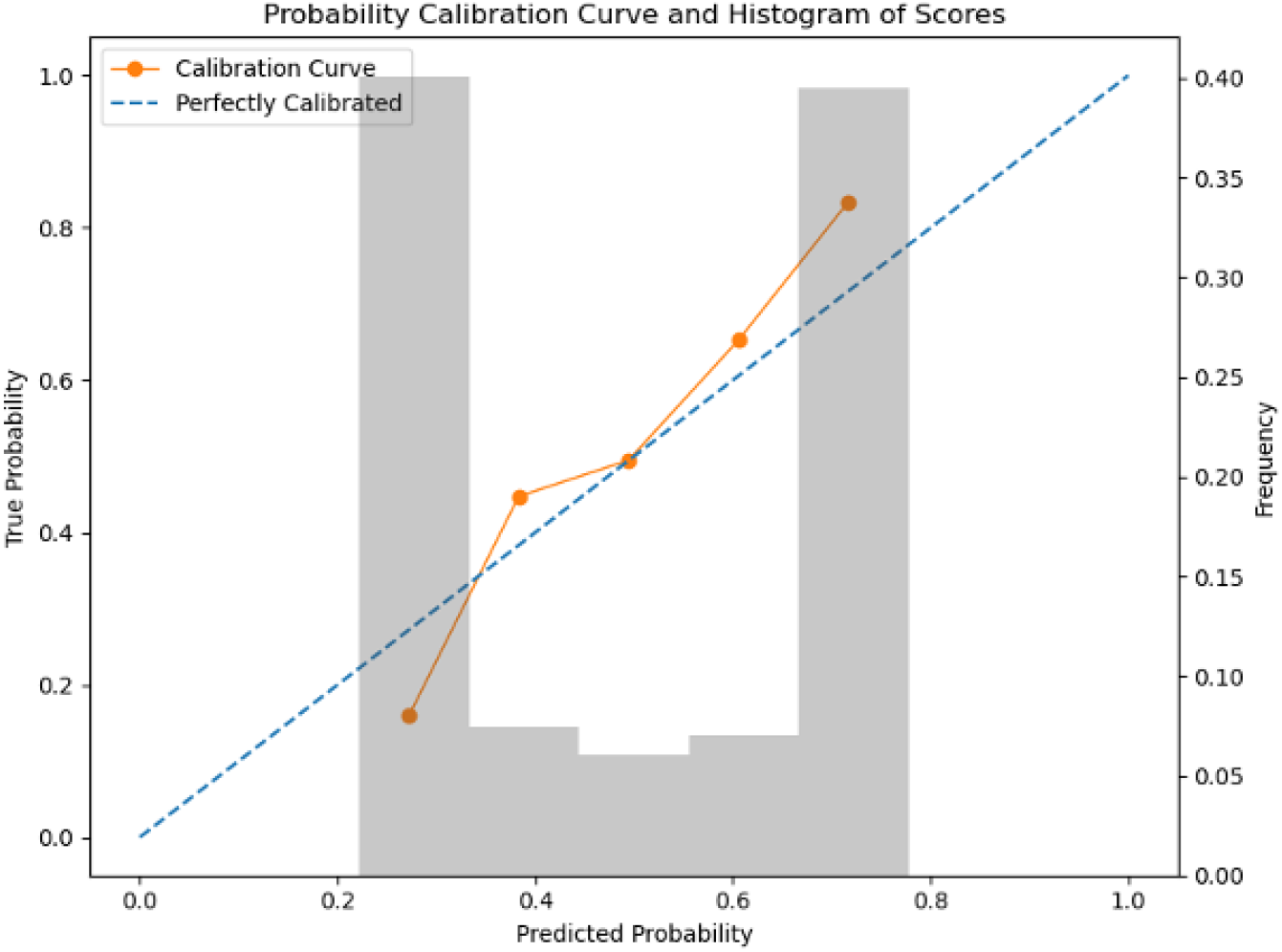
A calibration curve (in orange) for our trimodal model showing predicted probability on the x-axis and true probability on the left y-axis. Overlayed with the calibration curve is the prediction distribution (in gray). The distribution shows the prediction frequency, on the right y-axis, at each probability decile, the x-axis.

## Discussion

In this study, we examined the feasibility of multimodal deep learning for ARDS detection on heterogeneous datasets by training small models on top of larger pretrained models with different combinations of modalities. We found that small models built on top of the generalized embeddings of larger pretrained models can reliably classify ARDS in ICU patients. We also found that models trained on multiple modalities tend to outperform models trained on subsets of those modalities. Specifically, we showed that trimodal models tended to outperform bimodal models and that bimodal models tended to outperform unimodal models. We found all models to be generally well calibrated across the range of predicted probabilities.

Our findings build upon recent research [10, 9] that collectively supports the potential utility of machine learning to screen for ARDS. Our work extends previous work on machine learning-based ARDS detection in several ways. First, we tested the combinatorial addition of three clinical data sources, which to our knowledge, is the first trimodal approach to ARDS classification. Like other studies that examined one versus two data types, we observed incremental gains in performance with the use of two data sources [19, 20]. Of note, we found that the addition of either VWD or CXRs to EHR data achieved comparable performance. Images are ordered infrequently in clinical settings, whereas VWD is generated continuously, suggesting that the combination of EHR+VWD data could prove more suitable for frequent ML-based screening, especially for patients without ARDS when mechanical ventilation is initiated [21, 9], and points to the potential utility of extending continuous physiologic monitoring signals to multimodal deep learning [22, 23, 24, 25].

Our finding of additional, albeit modest, gains in performance with the use of three data sources suggests that the use of distinct but complementary data types may enable more effective classifiers to be developed. While our work demonstrates the utility of adding a third, more complex modality to an ARDS detection model and provides the flexibility to add more, our results highlight the need for additional research into other difficult-to-diagnose conditions and those with multimodal diagnostic criteria. Additional research could also help determine whether the added complexity and computational costs of multimodal model development are consistently beneficial, and whether certain conditions benefit more or less from the inclusion of certain data types.

We were surprised by our analysis of model performance in subgroups of “pure” ARDS (Group 1) versus patients with ARDS and concurrent chronic lung disease, congestive heart failure, or severe obesity (Group 2). We hypothesized that we would see a relatively larger improvement in measures of discrimination with the bimodal and trimodal models in Group 2 due to the greater medical complexity of that subgroup, where the addition of additional information content might help models to learn ARDS from non-ARDS cases. While bimodal and trimodal models saw consistent, and in many cases substantial, improvements in performance in Group 1 patients, Group 2 patients did not see consistent gains with additional data types. Our findings, if replicated, suggest that multimodal classification of conditions like ARDS have gaps. This is likely due to the known imperfect validity of ARDS diagnostic criteria which have undergone a continuous degree of debate throughout the last number of decades [26, 27]. Thus, the substantial physiologic overlap between clear and ambiguous cases [28] may not simply be addressed with a “more is better” strategy and instead may require subgroup-specific approaches to model development. Validation in multiple clinical centers would shed light on the utility of the “more is better” approach and would help determine if our results are an artifact of the three modalities chosen and/or local practices.

Our work also extends prior research in how it used pretrained models. Our approach used two pretrained models, one for CXR data and one for VWD, and then trained smaller models on top of the pretrained models, which are frozen during training. In the past, ARDS models have employed training strategies like pretraining a model on a general CXR dataset and finetuning it on a smaller ARDS dataset to make unimodal predictions [6], or using pretrained models to extract features that are then combined with tabular features to train traditional ML models [29, 30, 31, 9]. The most similar work to ours trains only the final layers of a pretrained model on ARDS-specific data [8]. However, this work built their multimodal model by ensembling the predictions of unimodal models, while our modalities are combined into a single model. Moreover, we could not find any pre-existing ARDS detection work that uses pretrained models for waveform data.

Our use of pretrained models also allowed us to extend prior research in how many patients are used during training. Our work used only 220 patients, while the smallest training set used in any of the work we found was >1.2k patients. Our observed performance in the trimodal model, which is similar to other published studies despite our small sample size [26, 20], suggests that using distinct but complimentary data types may be able to compensate for a relatively small learning space, which is common in biomedical research settings. Further, the level of performance reached on such a small sample space suggests a potential for improved performance in larger datasets if they are available.

### Technical Innovation and Broader Impact

Our approach serves as a proof of concept for adapting ideas from the broader world of ML and multimodal ML research. Using a small classification network on top of a larger, frozen pretrained network has become common practice for image classification [32]. The concept of using a projection network to reduce the dimensionality of a feature space is common for networks like autoencoders [33], and bottlenecks have also been used for multimodal networks outside of the biomedical space [34]. By combining many of these ideas, we showed the viability of multimodal deep learning on small heterogeneous datasets. Deep learning is notoriously data-hungry, and our application of pretrained models, combined with carefully selected architectures, demonstrated the feasibility of developing accurate, well-calibrated models, even with limited training data.

Our approach to handling multimodal data could serve as a template for other clinical applications where multiple data sources provide complementary information. The architecture we developed, with its modality-specific encoders and joint classification head, could be adapted for other conditions where diagnosis relies on the integration of multiple data types, and could easily be extended to incorporate additional modalities, such as other imaging modalities, medication data, or even clinical notes. This is particularly relevant in critical care, where clinicians routinely integrate data from multiple sources to make complex decisions. Moreover, the framework proposed in this paper is particularly well suited for small sample size multimodal datasets. The latter fact makes our framework especially valuable in clinical environments where data scarcity is often a barrier to applying advanced ML techniques.

### Limitations

Our study has several important limitations. First, the single-center nature of our study limits our ability to assess generalizability across different clinical settings and patient populations. ARDS, by definition, occurs across a wide spectrum of inciting conditions, and thus, our ARDS patient population may not be generalizable to all ARDS cases. Different institutions may have different ventilator management strategies, although our institution follows ARDSnet protocols. ARDS patients may also have been imaged at different intervals. These factors could affect the relative contributions of each data type to multimodal model classification performance. Second, while our sample size was sufficient to demonstrate the potential of our approach, a larger dataset would be needed to fully validate the model’s performance and ensure robust learning of complex cross-modal patterns. This is particularly important for the more complex ARDS cases in Group 2, where more examples would help better understand the relative contributions of each data type to model performance in less clear-cut cases and in key ARDS subgroups [35, 36].

Third, our retrospective study design was chosen to allow investigation of the effects of multimodal deep learning on ARDS classification but doesn’t address several challenges that would arise in real-world implementation. These include the need for real-time data processing, handling of missing or delayed data, and integration with clinical workflows. Because ARDS remains challenging to diagnose even after 24 hours of meeting diagnostic criteria[1], models focused on classification after this 24-hour window may still have utility. However, early recognition and frequent reassessment would facilitate the use of evidence-based therapies in conditions like ARDS where earlier treatment is thought to improve survival [3].

Finally, while our models showed good calibration within their ranges of predicted values, the absence of predictions in certain probability ranges (0.0-0.2 and 0.8-1.0) suggests some limitations in the models’ ability to express very high or very low confidence. This could be addressed in future work through improved model architecture or training approaches, including the potential inclusion of clinician uncertainty regarding the target label into the training process [37].

## Conclusion

Our results demonstrate the potential of multimodal deep learning to progressively improve ARDS classification by increasing the number of data types. Moreover, our methodological approach was able to produce performant models using small training datasets. However, we found heterogeneous performance with different combinations of modalities and in subgroups of ARDS patients, highlighting that more may not always be better. By showing that the integration of multiple data modalities can improve diagnostic performance, even in ambiguous cases, our work contributes to the broader goal of developing AI systems that can meaningfully support clinical decision-making in complex conditions and high-stakes clinical environments.

## Data Availability

Data for this study are not publicly available.

## Acknowledgments

The authors acknowledge support from NIH grant R01HL16351. The authors also acknowledge Anna Liu and Irene Cortes-Puch, MD for their work on data preparation, and the UC Davis Health Information Technology Data Provisioning Core and Health Computing Core.

## Supplementary Materials

### Model Training

We optimized the following hyperparameters during training:

- *D*_*e*_: 8, 16, or 32 dimensions
- *Proj*(·) size: one- or two-layer network
- Dropout rate in *Proj*(·) and *Class*(·): 50%, 70%, or 90%

Inspired by works using a contrastive pretraining step [11, 32], we also tried a version of our model where the projection layers are jointly trained to maximize the cosine similarity of modality-specific embeddings of the same patient. However, performance on the validation set was generally better using the approach described in the paper, so this contrastive approach was not explored further.

A full list of numerical and categorical features in EHR-derived tabular data is shown in Supplementary Table 4. A visual depiction of the splitting procedure used during training can be seen in Supplementary Figure 3.

**Table 4.** Full list of numerical and categorical features in EHR data.

| Numerical Features | Categorical Features |
| --- | --- |
| total_intake_ml | smoking_usage |
| total_output_ml | tobacco_user |
| sf | age_at_admsn_yr |
| sf97 | sex |
| osi | race |
| osi97 | ethnicity |
| temperature_celsius | adi_state_score |
| respiratory_rate | ca_hpi_percentile |
| heart_rate | ca_hpi_quartile |
| fio2 | elixhauser_weighted_score |
| spo2 | elixhauser_comorbidity_count |
| lab_pf_ratio_res |  |
| lab_white_blood_cell_res |  |
| lab_neutrophil_abs_manual_res |  |
| lab_neutrophil_abs_auto_res |  |
| lab_bands_percent_res |  |
| lab_hemoglobin_res |  |
| lab_platelet_res |  |
| lab_inr_res |  |
| lab_sodium_res |  |
| lab_potassium_res |  |
| lab_chloride_res |  |
| lab_bicarbonate_res |  |
| lab_e_gfr_res |  |
| lab_serum_glucose_res |  |
| lab_bilirubin_total_res |  |
| lab_asparate_transminase_res |  |
| lab_alanine_transferase_res |  |
| lab_alkaline_phosphatase_res |  |
| lab_lipase_res |  |
| lab_bnp_res |  |
| lab_creatinine_res |  |
| lab_bun_res |  |
| bp_map_cuff |  |
| bp_cuff_systolic |  |
| bp_cuff_diastolic |  |
| bp_map_a_line |  |
| bp_a_line_systolic |  |
| bp_a_line_diastolic |  |

**Table 5.** The statistical significance between average AUROC, as measured by a two-sample t-test, between models trained on different modalities. P-values are rounded to three decimal places.

| Modality 1 | Modality 2 | P-value |
| --- | --- | --- |
| Tabular | VWD+Tabular | 0.538 |
| Tabular | X-ray+Tabular | 0.166 |
| Tabular | Trimodal | 0.005 |
| X-ray | VWD+X-ray | 0.000 |
| X-ray | X-ray+Tabular | 0.000 |
| X-ray | Trimodal | 0.000 |
| VWD | VWD+Tabular | 0.000 |
| VWD | VWD+X-ray | 0.000 |
| VWD | Trimodal | 0.000 |
| VWD+Tabular | Trimodal | 0.039 |
| VWD+X-ray | Trimodal | 0.000 |
| X-ray+Tabular | Trimodal | 0.179 |

**Table 6.**
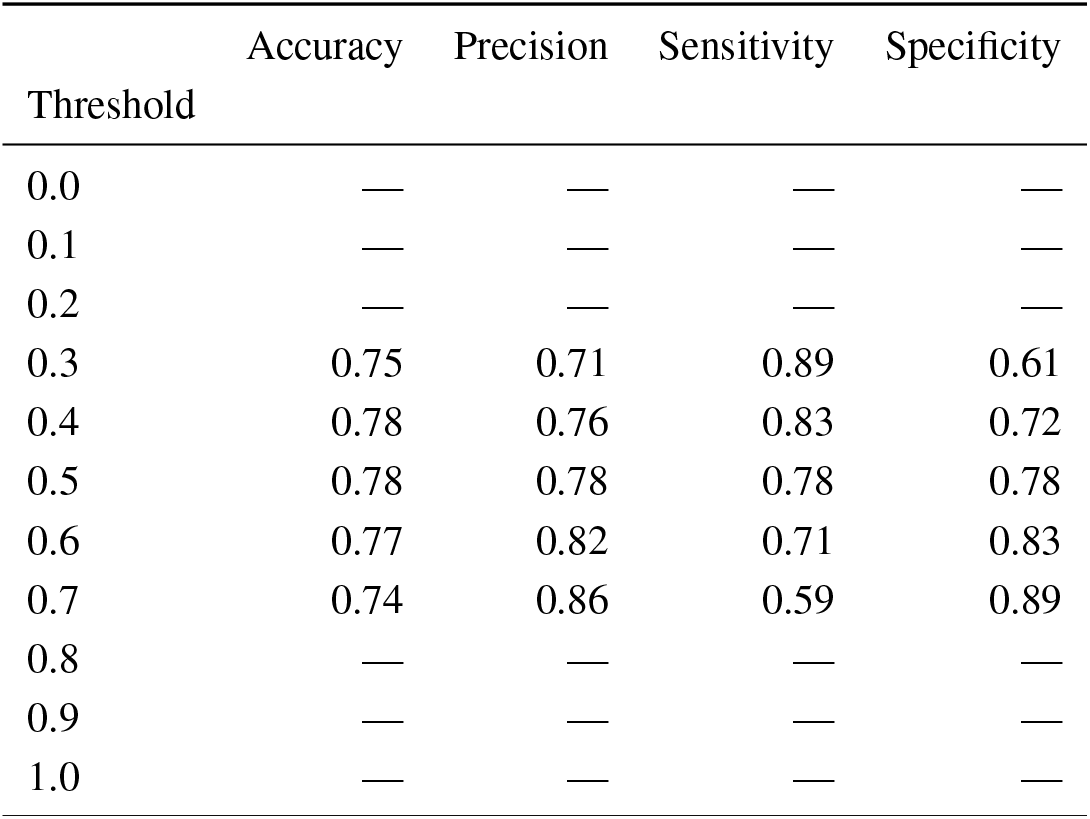
Average trimodal model performance characteristics across predicted probability thresholds.

| Threshold | Accuracy | Precision | Sensitivity | Specificity |
| --- | --- | --- | --- | --- |
| 0.0 | — | — | — | — |
| 0.1 | — | — | — | — |
| 0.2 | — | — | — | — |
| 0.3 | 0.75 | 0.71 | 0.89 | 0.61 |
| 0.4 | 0.78 | 0.76 | 0.83 | 0.72 |
| 0.5 | 0.78 | 0.78 | 0.78 | 0.78 |
| 0.6 | 0.77 | 0.82 | 0.71 | 0.83 |
| 0.7 | 0.74 | 0.86 | 0.59 | 0.89 |
| 0.8 | — | — | — | — |
| 0.9 | — | — | — | — |
| 1.0 | — | — | — | — |

### Statistical Tests

Two-sample t-tests were performed between the average AUROC value for every modality combination. They are in Supplementary Table 5.

### Model Calibration

In addition to, shows some confusion matrix statistics stratified by prediction threshold for the trimodal model. As noted in the paper, the models didn’t make any predictions <0.2 and <0.8.

In addition to Figure 2, Supplementary Table 6 shows some confusion matrix statistics stratified by prediction threshold for the trimodal model. As noted in the paper, the models didn’t make any predictions <0.2 and <0.8 Further, calibration curves for all models can be seen in.

Furthermore, Supplementary Figure 4 shows the same plot as Figure 2 for all modality combinations. Supplementary Figure 5 shows the distribution of holdout AUROC values for each tested modality combination.

**Figure 3.**
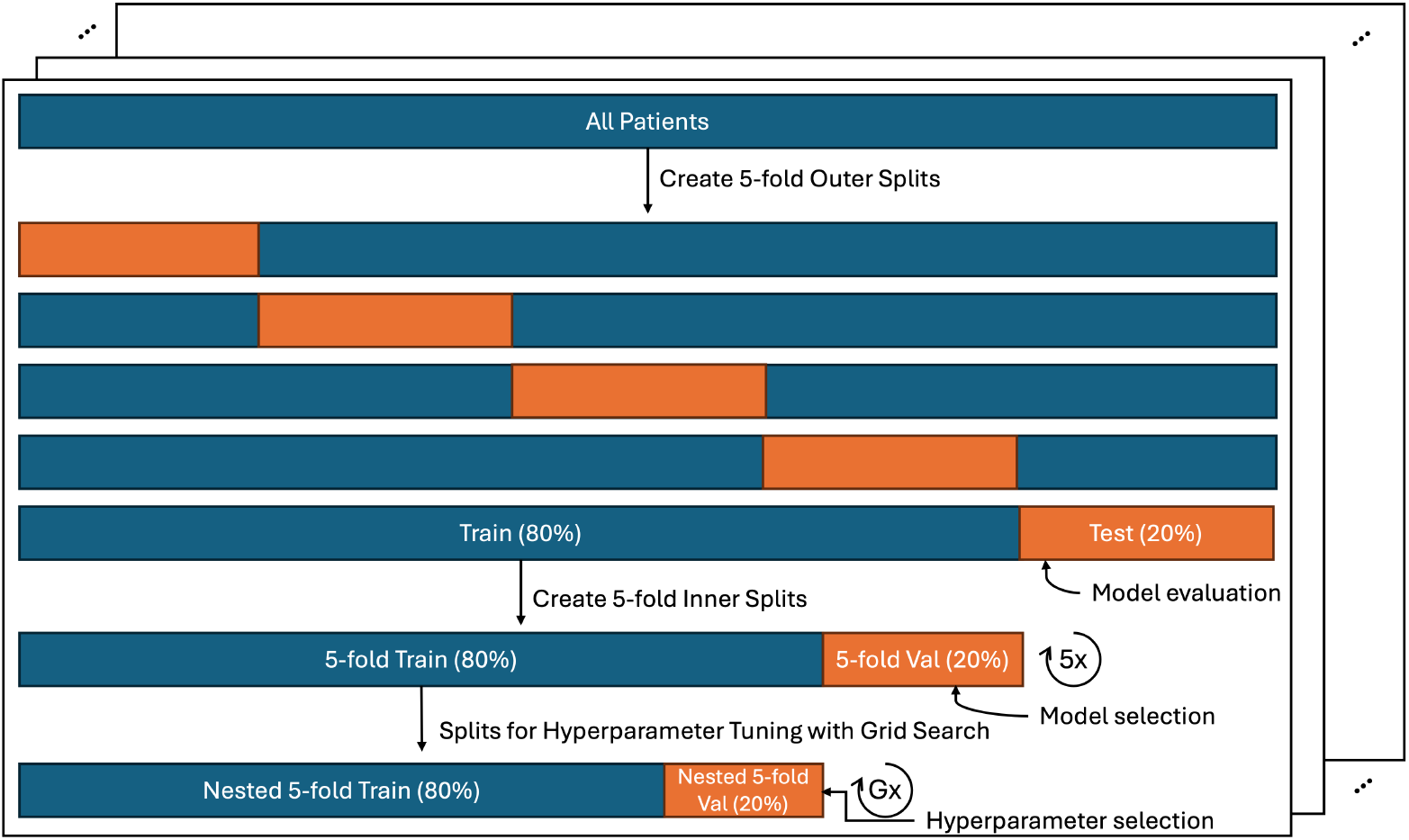
Splitting procedure for experimentation. Outer splits are created as a 5-fold split of the full patient dataset. Inner fold splits are created as a 5-fold split of each outer fold train split. Hyperparameter tuning was conducted by randomly splitting each inner fold train split G times, where G represents the number of hyperparameter combinations in a grid search. Final model performance was assessed on the test split of the outer folds. The full procedure was repeated 10 times.

**Figure 4.**
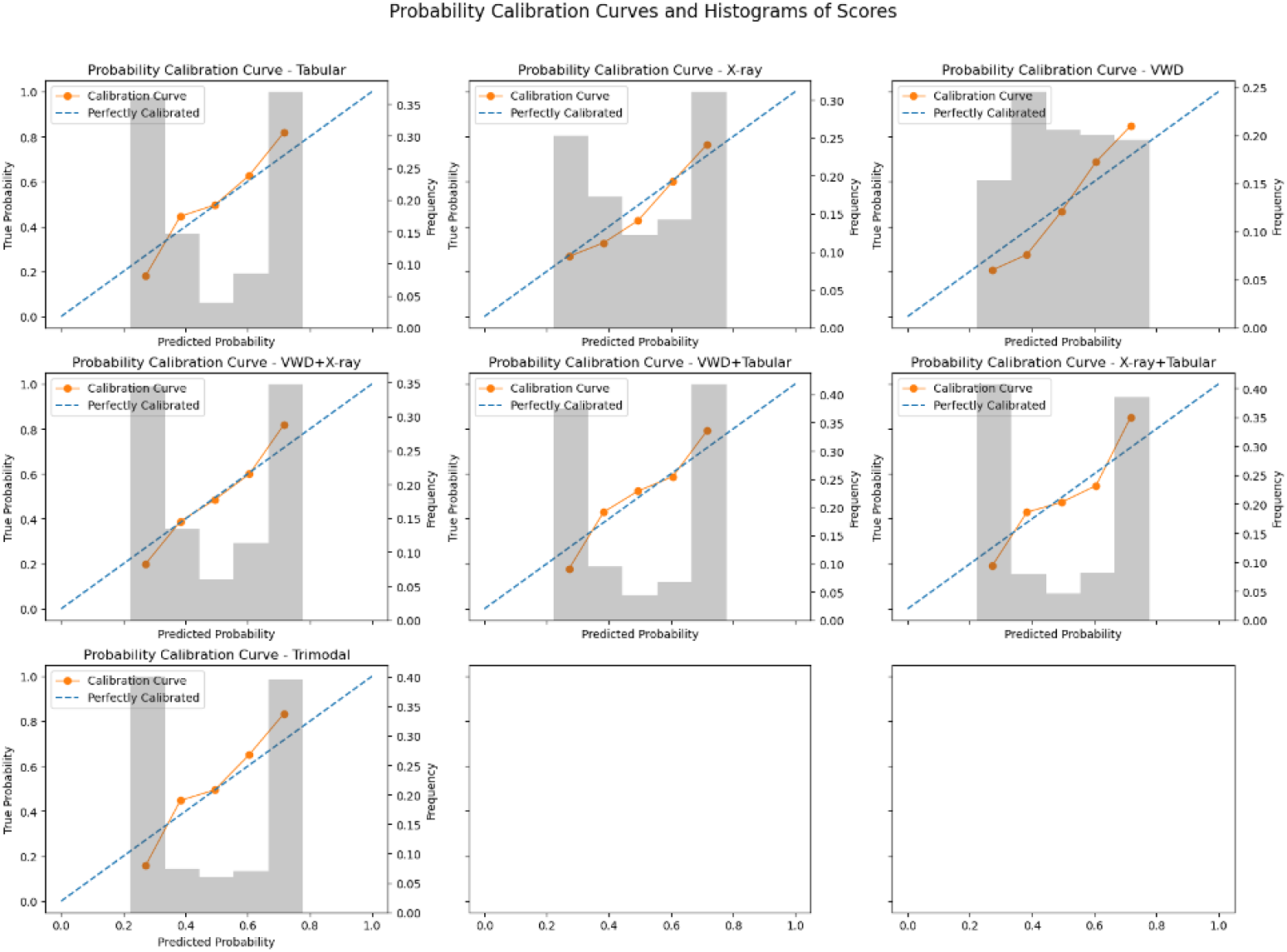
Calibration plots for models trained on all modality combinations.

**Figure 5.**
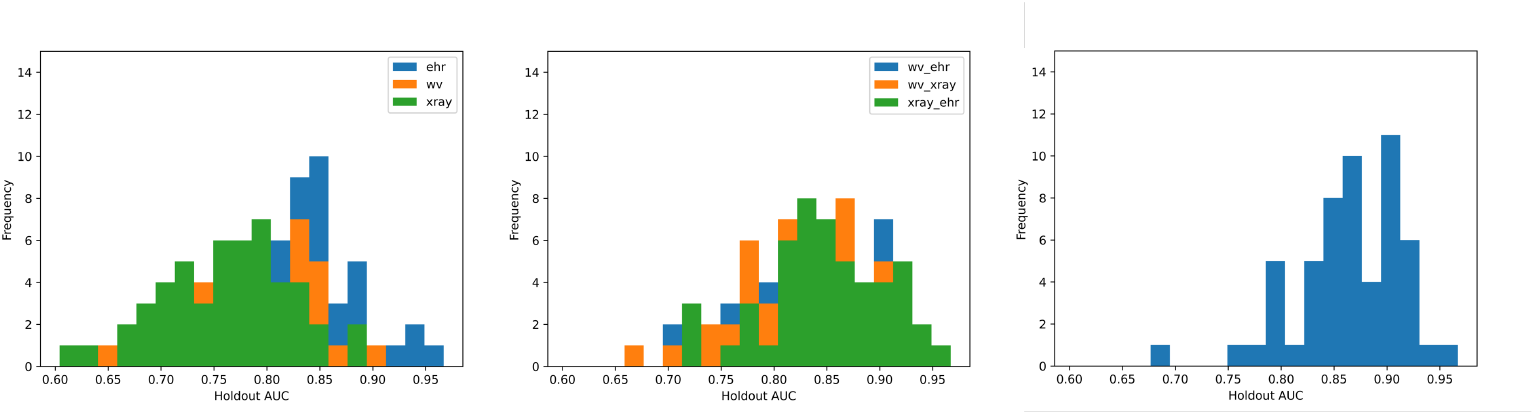
AUROC distributions for unimodal (left), bimodal (center), and trimodal (right) models.

## Footnotes

1 for first 24 hours post first ICU admission

## References

[1] G. Bellani et al. “Epidemiology, patterns of care, and mortality for patients with acute respiratory distress syndrome in intensive care units in 50 countries”. In: Jama 315 (2016), pp. 788–800.

[2] M. Matthay et al. “Acute respiratory distress syndrome”. In: Nature Reviews Disease Primers 5 (2019), p. 18.

[3] Dale M Needham et al. “Timing of low tidal volume ventilation and intensive care unit mortality in acute respiratory distress syndrome. A prospective cohort study”. In: American journal of respiratory and critical care medicine 191.2 (Jan. 2015). [Online; accessed 2025-07-07], pp. 177–85. ISSN: 1073-449X. DOI: 10.1164/rccm.201409-1598OC.

[4] C. Weiss et al. “Low tidal volume ventilation use in acute respiratory distress syndrome”. In: Critical Care Medicine 44 (2016), pp. 1515–1522.

[5] ARDS Definition Task Force et al. “Acute respiratory distress syndrome: the Berlin Definition”. In: JAMA 307.23 (June 2012). [Online; accessed 2025-07-07], pp. 2526–33. ISSN: 0098-7484. DOI: 10.1001/jama.2012.5669.

[6] M. Sjoding et al. “Deep learning to detect acute respiratory distress syndrome on chest radiographs: a retrospective study with external validation”. In: The Lancet Digital Health 3 (). Publisher: Elsevier. (2021,6), e340–e348. URL: https://www.thelancet.com/journals/landig/article/PIIS2589-7500(21)00056-X/fulltext.

[7] G. Rehm et al. “Use of machine learning to screen for acute respiratory distress syndrome using raw ventilator waveform data”. In: Critical Care Explorations 3 (2021), e0313.

[8] K. Pai et al. “Artificial intelligence–aided diagnosis model for acute respiratory distress syndrome combining clinical data and chest radiographs”. In: Digital Health 8 (). p 76221120317 (2022,8), p. 205520. URL: https://www.ncbi.nlm.nih.gov/pmc/articles/PMC9386858/.

[9] E. Levy et al. “Development and External Validation of a Detection Model to Retrospectively Identify Patients With Acute Respiratory Distress Syndrome”. In: Critical Care Medicine (2025), pp. 10–1097.

[10] Francesca Rubulotta et al. “Machine Learning Tools for Acute Respiratory Distress Syndrome Detection and Prediction”. In: Critical Care Medicine 52.11 (2024), pp. 1768–1780.

[11] Y. Zhang et al. “Contrastive Learning of Medical Visual Representations from Paired Images and Text”. [cs]. arXiv, 9). 2022. 2010.00747. URL: http://arxiv.org/abs/2010.00747.

[12] V. Lopez et al. “An insight into classification with imbalanced data: Empirical results and current trends on using data intrinsic characteristics”. In: Information Sciences 250 (2013), pp. 113–141.

[13] F. Schmid, K. Koutini, and G. Widmer. “Efficient large-scale audio tagging via transformer-to-cnn knowledge distillation”. In: ICASSP 2023-2023 IEEE International Conference On Acoustics, Speech And Signal Processing (ICASSP). 2023, pp. 1–5.

[14] F. Schmid, K. Koutini, and G. Widmer. “Low-complexity audio embedding extractors”. In: 2023 31st European Signal Processing Conference (EUSIPCO). 2023, pp. 451–455.

[15] B. Boecking et al. “Making the Most of Text Semantics to Improve Biomedical Vision–Language Processing”. [cs]. 2022. 2204.09817. URL: http://arxiv.org/abs/2204.09817.

[16] K. He et al. “Deep residual learning for image recognition”. In: Proceedings Of The IEEE Conference On Computer Vision And Pattern Recognition. 2016, pp. 770–778.

[17] J. Yoon et al. “Vime: Extending the success of self-and semi-supervised learning to tabular domain”. In: Advances In Neural Information Processing Systems 33 (2020), pp. 11033–11043.

[18] Pauli Virtanen et al. “SciPy 1.0: Fundamental Algorithms for Scientific Computing in Python”. In: Nature Methods 17 (2020), pp. 261–272. DOI: 10.1038/s41592-019-0686-2.

[19] Sarah Jabbour et al. “Combining chest X-rays and electronic health record (EHR) data using machine learning to diagnose acute respiratory failure”. In: Journal of the American Medical Informatics Association : JAMIA 29.6 (May 2022). [Online; accessed 2025-07-07], pp. 1060–1068. ISSN: 1067-5027. DOI: 10.1093/jamia/ocac030.

[20] Yaxin Xiong et al. “Accuracy of artificial intelligence algorithms in predicting acute respiratory distress syndrome: a systematic review and meta-analysis”. In: BMC medical informatics and decision making 25.1 (Jan. 2025). [Online; accessed 2025-07-07], p. 44. ISSN: 1472-6947. DOI: 10.1186/s12911-025-02869-0.

[21] Daniel Zeiberg et al. “Machine learning for patient risk stratification for acute respiratory distress syndrome”. In: PloS one 14.3 (Mar. 2019). [Online; accessed 2025-07-07], e0214465. ISSN: 1932-6203. DOI: 10.1371/journal.pone.0214465.

[22] Wei-Long Zheng et al. “Predicting neurological outcome in comatose patients after cardiac arrest with multiscale deep neural networks”. In: Resuscitation 169 (Dec. 2021). [Online; accessed 2025-07-11], pp. 86–94. ISSN: 0300-9572. DOI: 10.1016/j.resuscitation.2021.10.034.

[23] Awni Y. Hannun et al. “Cardiologist-level arrhythmia detection and classification in ambulatory electrocardiograms using a deep neural network”. In: Nature Medicine 25.1 (Jan. 2019), pp. 65–69. ISSN: 1078-8956. DOI: 10.1038/s41591-018-0268-3. URL: http://dx.doi.org/10.1038/s41591-018-0268-3.

[24] Stefan Gustafsson et al. “Development and validation of deep learning ECG-based prediction of myocardial infarction in emergency department patients”. In: Scientific Reports 12.1 (Nov. 2022). ISSN: 2045-2322. DOI: 10.1038/s41598-022-24254-x. URL: http://dx.doi.org/10.1038/s41598-022-24254-x.

[25] Marleen C Tjepkema-Cloostermans et al. “Outcome Prediction in Postanoxic Coma With Deep Learning”. In: Critical care medicine 47.10 (Oct. 2019). [Online; accessed 2025-07-12], pp. 1424–1432. ISSN: 0090-3493. DOI: 10.1097/CCM.0000000000003854.

[26] Arnaud W Thille et al. “Comparison of the Berlin definition for acute respiratory distress syndrome with autopsy”. In: American journal of respiratory and critical care medicine 187.7 (Apr. 2013). [Online; accessed 2025-07-07], pp. 761–7. ISSN: 1073-449X. DOI: 10.1164/rccm.201211-1981OC.

[27] Niall D Ferguson et al. “Screening of ARDS patients using standardized ventilator settings: influence on enrollment in a clinical trial”. In: Intensive care medicine 30.6 (June 2004). [Online; accessed 2025-07-07], pp. 1111–6. ISSN: 0342-4642. DOI: 10.1007/s00134-004-2163-2.

[28] Michael W Sjoding et al. “Differences between Patients in Whom Physicians Agree and Disagree about the Diagnosis of Acute Respiratory Distress Syndrome”. In: Annals of the American Thoracic Society 16.2 (Feb. 2019). [Online; accessed 2025-07-07], pp. 258–264. ISSN: 2325-6621. DOI: 10.1513/AnnalsATS.201806-434OC.

[29] N. Reamaroon et al. “Automated detection of acute respiratory distress syndrome from chest X-Rays using Directionality Measure and deep learning features”. In: Computers In Biology And Medicine 134 (2021), p. 104463.

[30] B. Afshin-Pour et al. “Discriminating Acute Respiratory Distress Syndrome from other forms of respiratory failure via iterative machine learning”. In: Intelligence-Based Medicine 7 (2023), p. 100087.

[31] A. Gandomi et al. “ARDSFlag: an NLP/machine learning algorithm to visualize and detect high-probability ARDS admissions independent of provider recognition and billing codes”. In: BMC Medical Informatics And Decision Making 24 (2024), p. 195.

[32] T. Chen et al. “A simple framework for contrastive learning of visual representations”. In: International Conference On Machine Learning. 2020, pp. 1597–1607.

[33] Jürgen Schmidhuber. “Deep learning in neural networks: An overview”. In: Neural networks 61 (2015), pp. 85–117.

[34] Arsha Nagrani et al. “Attention bottlenecks for multimodal fusion”. In: Advances in neural information processing systems 34 (2021), pp. 14200–14213.

[35] Carolyn S Calfee et al. “Subphenotypes in acute respiratory distress syndrome: latent class analysis of data from two randomised controlled trials”. In: The Lancet. Respiratory medicine 2.8 (Aug. 2014). [Online; accessed 2025-07-07], pp. 611–20. ISSN: 2213-2600. DOI: 10.1016/S2213-2600(14)70097-9.

[36] Patricia L Valda Toro et al. “Rapidly improving ARDS differs clinically and biologically from persistent ARDS”. In: Critical care (London, England) 28.1 (Apr. 2024). [Online; accessed 2025-07-07], p. 132. ISSN: 1364-8535. DOI: 10.1186/s13054-024-04883-6.

[37] Narathip Reamaroon et al. “Accounting for Label Uncertainty in Machine Learning for Detection of Acute Respiratory Distress Syndrome”. In: IEEE journal of biomedical and health informatics 23.1 (Jan. 2019). [Online; accessed 2025-07-07], pp. 407–415. ISSN: 2168-2194. DOI: 10.1109/JBHI.2018.2810820.

